# Amputee, clinician, and regulator perspectives on current and prospective upper extremity prosthetic technologies

**DOI:** 10.1101/2020.08.07.20170209

**Authors:** Julie S. Rekant, Lee E. Fisher, Michael L. Boninger, Robert A. Gaunt, Jennifer L. Collinger

## Abstract

Existing prosthetic technologies for people with upper limb amputation are being adopted at moderate rates and unfortunately these devices are often abandoned. The aims of this study were to: 1) understand the current state of satisfaction with upper extremity prostheses, 2) solicit feedback about prosthetic technology and important device design criteria from amputees, clinicians, and device regulators, and 3) compare and contrast these perspectives to identify common or divergent priorities. Twenty-one adults with upper limb loss, 35 clinicians, and 3 regulators completed a survey on existing prosthetic technologies and a conceptual sensorimotor prosthesis driven by implanted myoelectric electrodes with sensory feedback provided via stimulation of dorsal root ganglion. User and clinician ratings of satisfaction with existing prosthetic devices were similar. While amputees, clinicians, and regulators were similarly accepting of technology in general, amputees were most accepting of the proposed implantable sensorimotor prosthesis. Overall, stakeholders valued user-centred outcomes such as individualized task goals, improved quality of life, device reliability, and user safety; a large emphasis was put on these last two outcomes by regulators. The results of this study provide insight into the priorities of amputees, clinicians, and regulators that will inform future upper-limb prosthetic design and clinical trial protocol development.

## Introduction

Approximately 2 million Americans are living with limb loss, and of these, 20% affect the upper limb ^[1, 2]^. Prosthesis adoption among upper-limb amputees is considerably less than among lower-limb amputees, and, once adopted, upper extremity prosthetic devices are abandoned nearly half the time ^[3-7]^. This suggests that existing upper limb prosthetic technology is either not accessible to those who need it, not meeting the needs of its users, or both ^[8]^.

Prosthesis users are a heterogeneous group of people and therefore devices seek to meet user needs in a variety of ways. For many, an upper extremity prosthesis is an aesthetic placeholder for a missing limb, while for others it is intended to be a more functional replacement. Current upper extremity prosthetic technology varies from cosmetic devices with minimal manipulation abilities but naturalistic appearance, to advanced robotic arms that articulate in multiple directions at many joints, allowing users to control wrist, hand, and finger movements ^[9-11]^. Though more advanced powered prostheses are coming to market ^[12, 13]^, comprehensive studies of user satisfaction and adoption of these devices are limited, with nearly all studies specifically considering the DEKA Arm ^[11, 14-18]^. Resnik, et al., evaluated the DEKA Arm and iterated on its design based on user feedback, ultimately improving functionality, weight, and portability with successive generations of the device ^[11, 14, 16, 17]^. However, upper-extremity prosthesis users continue to report a preference towards their existing devices over these advanced robotic prostheses ^[16, 18]^.

Most commercial myoelectric upper extremity prostheses are limited to 1-2 controllable degrees of freedom using surface recordings of muscle activity (e.g. Ottobock DynamicArm and MyoHandsJ^19, 20]^, ArmDynamics Dynamic Arm Elbow ^[21]^). However, neural interfaces are being investigated to provide high fidelity command signals for more dexterous prostheses ^[12, 13, 16, 22]^ and to provide sensory feedback ^[23-33]^. The lack of sensory feedback has been identified as a drawback of current upper extremity prostheses ^[34-37]^. However, there is a paucity of research regarding stakeholder perspectives on these sensory restoration techniques. In a prosthesis user focus group, Zheng et al. observed mixed views on sensory-enabled devices with many participants expressing scepticism about theoretical or research-based technologies ^[38]^. A few studies have demonstrated performance improvements, especially for complex tasks, with the integration of sensory feedback ^[26-29, 39]^. Further, case studies have shown changes in psychosocial factors such as prosthetic embodiment ^[40, 41]^, quality of life, and self-efficacy in users of a sensory-enabled prosthesis ^[15, 39]^. It is important to understand not only how devices are functioning, but also how they are meeting the needs they are intended to address and where they continue to fall short.

The objectives of this study were to understand the current state of satisfaction with upper extremity prostheses, identify stakeholder opinions on new technology and important prosthetic design criteria as they relate to advanced upper extremity prostheses, and compare and contrast the above perspectives across amputees, clinicians, and device regulators. These three groups completed surveys on device use and satisfaction, design priorities, and technology acceptance. They were then introduced to a proposed implantable sensorimotor neuroprosthesis and feedback on this device was collected. Understanding the priorities and values of stakeholders can direct the design process to develop a device that addresses current gaps in upper extremity prosthetic technology and one that is more likely to be adopted by users ^[14, 35, 42, 43]^. Additionally, information gathered in this study from a small group of device regulators adds perspective about which aspects of advanced prosthetic devices may present challenges during the regulatory review process and may subsequently affect the cost and accessibility of advanced neuroprosthetic devices ^[44]^.

## Methods

### Study Participants

Three stakeholder groups were recruited to participate in this study, which was approved by the University of Pittsburgh Institutional Review Board and the Human Research Protection Office of the Army Research Laboratory. Participants were included if they were over the age of 18 and identified with one of the following categories: person with upper limb amputation or congenital limb loss, clinician who works with people with limb loss, or person who acts as a regulator for prosthetic technology. Participants from the ‘amputee group’ were recruited through research registries, local rehabilitation and prosthesis clinics, and through advertisements with local and national organizations of interest to people with upper limb amputation. Clinicians were recruited from the local healthcare system as well as through advertisements with professional prosthetics and orthotics organizations. Finally, the regulator group was recruited by contacting the Office of Device Evaluation at the Food and Drug Administration.

### Survey Design

The primary component of the study was a survey that was available in both online and paper versions. Twenty percent of amputees (n=4), and all clinicians and regulators completed the online version of the survey. The surveys for each stakeholder group were designed by the investigators with input from local prosthetists and included questions from existing metrics ^[45, 46]^ as well as novel questions related to technology acceptance and neuroprosthetic design criteria. Many of the questions were common across all groups, with additional questions related to the impact of having an amputation and current prosthetic utilization asked of the amputee group. The following sections were included in each survey:

#### Demographics

All participants were asked to report gender, age, ethnicity, and race. Additional information on geographic location, type of housing, highest level of education completed, employment status, disability status, and source of health insurance was collected from the amputee group. The amputee group was also asked to report side and level of amputation, handedness prior to and following amputation, time since amputation, and reason for amputation.

Clinicians and regulators reported years of practice with the amputee population and details regarding their professional background.

#### Prosthesis History and Use

Questions from the Trinity Amputation and Prosthesis Experience Scale - Revised (TAPES-R) ^[46, 47]^ and the Disabilities of the Arm, Shoulder, and Hand (DASH) ^[45, 47, 48]^ were included within the survey to obtain information related to functional level, device utilization, and device satisfaction. The amputee group completed all questions from the TAPES- R that were relevant to upper-limb amputees, with wording adjusted to focus answers on experience with a prosthetic device as opposed to experience with an amputation. Participants with amputation that did not use a prosthesis were asked to report the most significant reason they did not use a device, and what features a prosthesis would need to have before they would consider using one. Clinicians also completed one section of the TAPES-R that asks how satisfied they were with the color, shape, appearance, weight, usefulness, reliability, fit, and comfort of existing prosthetic devices; they were asked to complete the questions three times with either a body-powered, myoelectric, or cosmetic prosthesis in mind. In accordance with TAPES-R scoring, color, shape, and appearance scores were combined into an aesthetic satisfaction rating, presented as a percentage; the remaining items were used to calculate a functional satisfaction rating ^[49]^. Regulators did not complete this section of the survey.

Since many physical activity questions from the TAPES-R are more relevant to lower limb amputees (e.g. can you climb one flight of stairs), the DASH activity section (questions 1-21 from the DASH questionnaire) was used to quantify the impact of amputation on activities that involve the upper limb. The DASH was used in its original form and then, taking inspiration from the OPUS Upper Extremity Functional Status survey ^[50]^, participants were asked to report which arm (intact, amputated/prosthetic, either, or both) they used to complete each task (included in Appendix A).

This section also included two questions to assess prosthetic design goals. The amputee and clinician groups were provided a free-text response area to report three activities they would like (or believe their patients would like) to be able to perform better with an upper limb prosthesis. Both groups were also asked to rate the importance of seven functions in a hand and wrist prosthesis: flexion/extension of all fingers simultaneously, individual finger flexion/extension, thumb flexion/extension, thumb abduction/adduction, wrist supination/pronation, wrist flexion/extension, and wrist abduction/adduction. Scores ranged from one (“Not at all important”) to five (“Extremely important”).

#### Technology Acceptance

This section consisted of a three-part, 24-item questionnaire to evaluate general opinions on technology, personal attitudes towards technology, and factors in technology adoption preferences^[51, 52]^. Seventeen questions related to technology acceptance were scored from one to five, where higher scores indicate greater acceptance. A total score as a percentage out of 100 was calculated to represent how accepting of technology the individual was. The remaining seven questions gathered opinions regarding design criteria that were important when choosing to use a technology (e.g. cost or ease of use). The full Technology Acceptance questionnaire can be found in Appendix B.

#### Neuroprosthetic Technology Video

After completing the technology acceptance questionnaire, participants were shown a short video describing a proposed sensorimotor neuroprosthesis, which we referred to as the “MyoTouch” for brevity though it was made clear that this was not an existing device (Supplementary Video 1). The proposed neuroprosthesis used muscle activity recorded from implanted electrodes to control the prosthesis and provided sensory feedback through stimulation of electrodes implanted over the cervical dorsal root ganglia. Importantly, we chose to present a single hypothetical device where the control scheme relied on recording myoelectric signals from residual muscles in the forearm and therefore was most appropriate for transradial amputees. A similar myoelectric control approach could be used with more proximal residual muscles. Similarly, stimulation of other locations in the peripheral or central nervous system could be used to provide sensory feedback.

#### Prospective Technology

Participants were asked about design characteristics and opinions towards an advanced sensorimotor neuroprosthesis such as the one described in the video (See Appendix C). First, subjects completed free response questions regarding their “likes” and “dislikes” about the device. Second, subjects were asked whether they agreed with statements related to the acceptance of the device where scores were ranked from 1 (strongly disagree) to 5 (strongly agree). A composite “neuroprosthesis acceptance score” was calculated for each participant using a similar method as is described for the “technology acceptance score” where a higher score reflects greater acceptance of the device and scores are presented as a percentage out of 100. Finally, participants were asked to select acceptable design characteristics for the proposed device (i.e. calibration time, reliability, and battery life). Regulators were asked additional questions about anticipated concerns related to regulatory approval and reimbursement.

### Data Analysis

All free text responses were thematically categorized by author J.R. and reviewed by a second author (J.C.) prior to analysis. Frequency of responses per category are reported throughout as proportions of total number of responses recorded per group. Scoring for DASH and TAPES-R questions followed previously reported methods ^[45, 46]^. All data were analyzed using SPSS Statistics Editor 24.0 (Armonk, NY). Normally-distributed continuous data were compared across groups using the one-way analysis of variance (ANOVA). Non-normally-distributed data were compared with the Mann-Whitney U Test (for two-group comparisons) or the Kruskal-Wallis H test (for comparison across all three groups). The relationship between demographics, TAPES-R scores, and MyoTouch acceptance scores were examined using the Spearman’s correlation. Significance was considered relative to an alpha value of 0.05.

## Results

### Demographics

Twenty-one amputees, 35 clinicians, and 3 regulators met the inclusion criteria and consented to participate in the study. Subject demographics are summarized in Table 1. Two amputees reported bilateral amputations: one had bilateral transradial amputations and one had bilateral transhumeral amputations. Of the 19 unilateral amputees included in this study, one had a partial hand amputation, one had a wrist amputation, eight had a transradial amputation, two had an elbow disarticulation, four had a transhumeral amputation, and three had a shoulder-level amputation. Eight participants had congenital limb loss and had a mean age of 36.2 ± 11.0 years. The other 13 amputee group participants had amputations secondary to trauma (n=9), infection (n=2), cancer (n=1), and compartment syndrome (n=1) and a mean time from amputation of 12.7 ± 15.5 years.

**Table 1.**
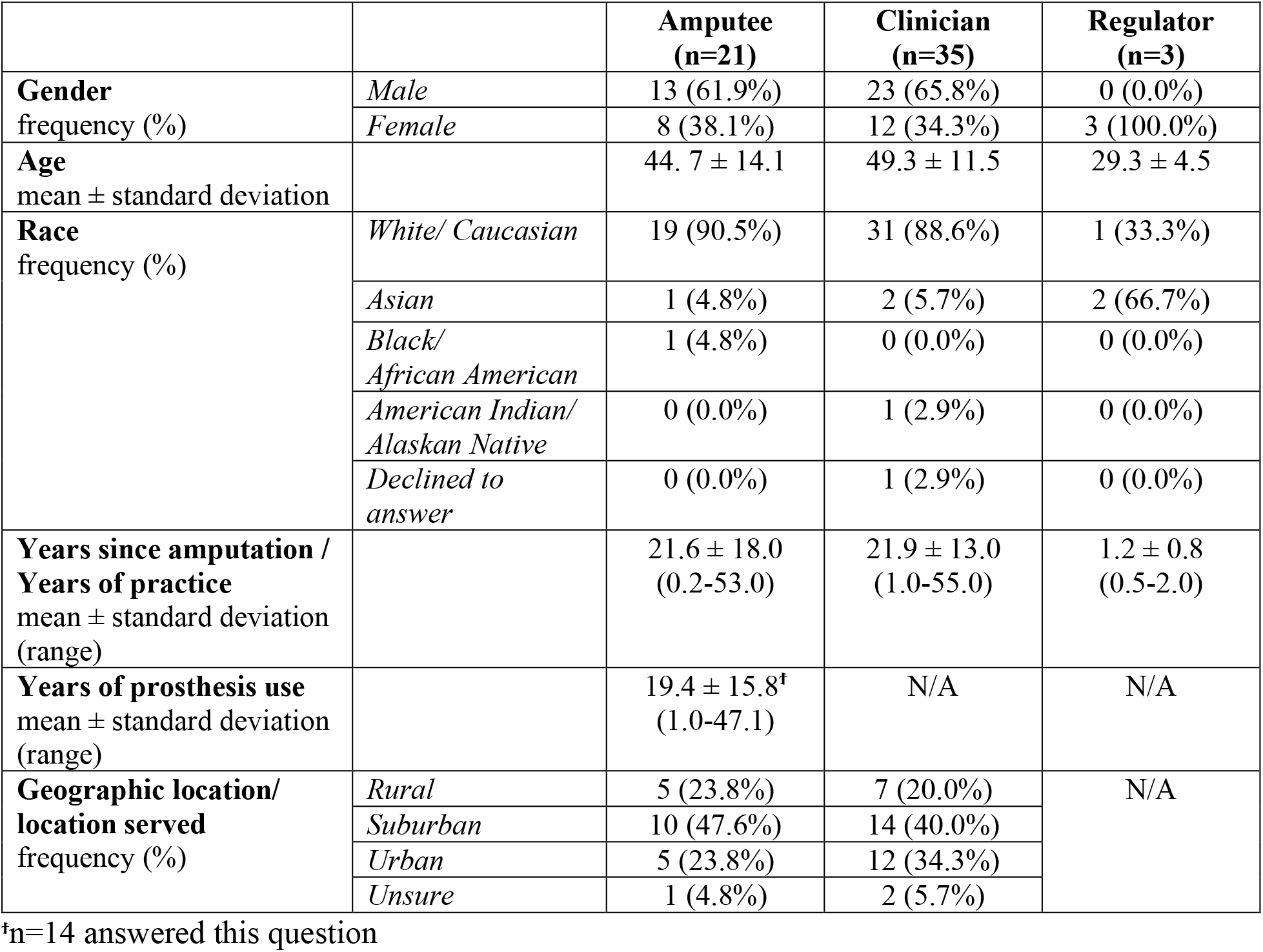
Subject demographics by group

All amputees reported living in a private residence (i.e. house, apartment, or condominium). Participants with amputations represented a wide variety of formal education levels, including 9^th^ through 11^th^ grade (n=1), high school diploma or GED (n=8), Associates Degree or Vocational/Technical School degree (n=5), Bachelor’s Degree (n=3), and Master’s Degree (n=4). Six participants were working full-time jobs at the time of the study, two were working part-time jobs, and 13 were not employed. Five individuals reported being on disability at the time of the study, while 12 felt their physical disability had decreased their income. Amputees reported receiving health insurance through Medicare or Medicaid (n=8), their past or present employer or their spouse or parents’ employer (n=10), a community-funded healthcare program (n=1), a combination of Medicare/Medicaid and an employer (n=1), and personally funded healthcare (n=1).

Clinicians involved in this study came from the professional backgrounds of Prosthetics (n=28), Occupational Therapy (n=3), Physical Therapy (n=2), Physiatry (n=1), and Neurosurgery (n=1).

All three of the regulators worked as medical device regulators at the FDA and reported two years or less of experience working with people with amputation.

#### Impact of amputation and history of prosthesis use

A majority of the amputee group reported having a prosthesis (17 of 21, 81%), indicating that this cohort is more active in terms of prosthesis use than the average group of upper limb amputees in the United States (55-65% prosthesis use) ^[5, 7]^. This may be because many upper limb amputees have a partial hand amputation (20%) ^[2, 5]^, whereas our cohort had more proximal amputations. Most amputees in this study reported primary use of a single device: body-powered (n=8), myoelectric (n=3), or cosmetic (n=2). However, three people reported approximately equal use of two devices: body-powered and myoelectric (n=2), myoelectric and body-powered/myoelectric hybrid (n=1), or myoelectric and a specialized function-specific adaptive device (n=1). One person reported approximately equal use of three devices (body-powered, myoelectric, and adaptive). Three of the four participants who did not have a device reported interest in obtaining a prosthesis but had not yet done so at the time of the survey. The fourth reported not having a prosthetic device because they “feel [they] can get through daily activities without needing one”. When prosthesis users chose not to use their devices, it was because the prosthesis was cumbersome or inefficient to use (n=5) or uncomfortable (n=2). Users and non-users primarily felt that prostheses needed to have better functionality (n=7) in order to be adopted. Other desired characteristics included being lightweight (n=2), comfortable (n=2), and having a life-like appearance (n=1).

The mean DASH score for the amputee group was 28.2% ± 18.9%, where a score of 0% indicates no disability and score of 100% indicates complete disability. The normative DASH score for similarly-aged healthy controls is 14% with a minimally clinically important difference (MCID) of 12.6% ^[53, 54]^. On average, the mean level of disability reported by amputees using the DASH (28.2%) exceeds the MCID compared to this normative value. This suggests that having an amputation has a meaningful and negative impact on function. In fact, all but one of the unilateral amputees elected to utilize their intact limb in some capacity in all tasks that they were able to accomplish, either independently or in conjunction with the involved arm. The activities from the DASH with the highest reported levels of difficulty were opening a jar, gardening, washing one’s back, cutting food, and participating in recreational activities.

TAPES-R adjustment scores were used to assess overall adjustment to prosthesis use among the amputee group. Scores can range from 1-4 with a higher score indicating better adjustment ^[49]^. The 16 individuals from the amputee group who responded to this section of the survey reported an average general adjustment of 3.2 ± 0.5, a social adjustment of 3.6 ± 0.5, and an adjustment to limitation of 2.5 ± 1.0. Responses to each question were examined to determine which aspects of adjustment were most difficult for the amputees. Six participants disagreed with the statement “I have adjusted to having a prosthesis” and 5 people disagreed with the statement “I have gotten used to wearing a prosthesis”. Prosthesis users felt most limited in the areas of work interference (n=9) and dependence on others (n=7).

TAPES-R satisfaction scores were evaluated for both the amputee and clinician groups to gain a better understanding of how well these groups feel current prosthetic devices are meeting the needs and expectations of users; users only rated devices they had personal experience with. Normalized satisfaction ratings from clinicians and amputees are shown in Figure 1 by device type. No significant differences in satisfaction ratings were observed between subject groups.

**Figure 1.**
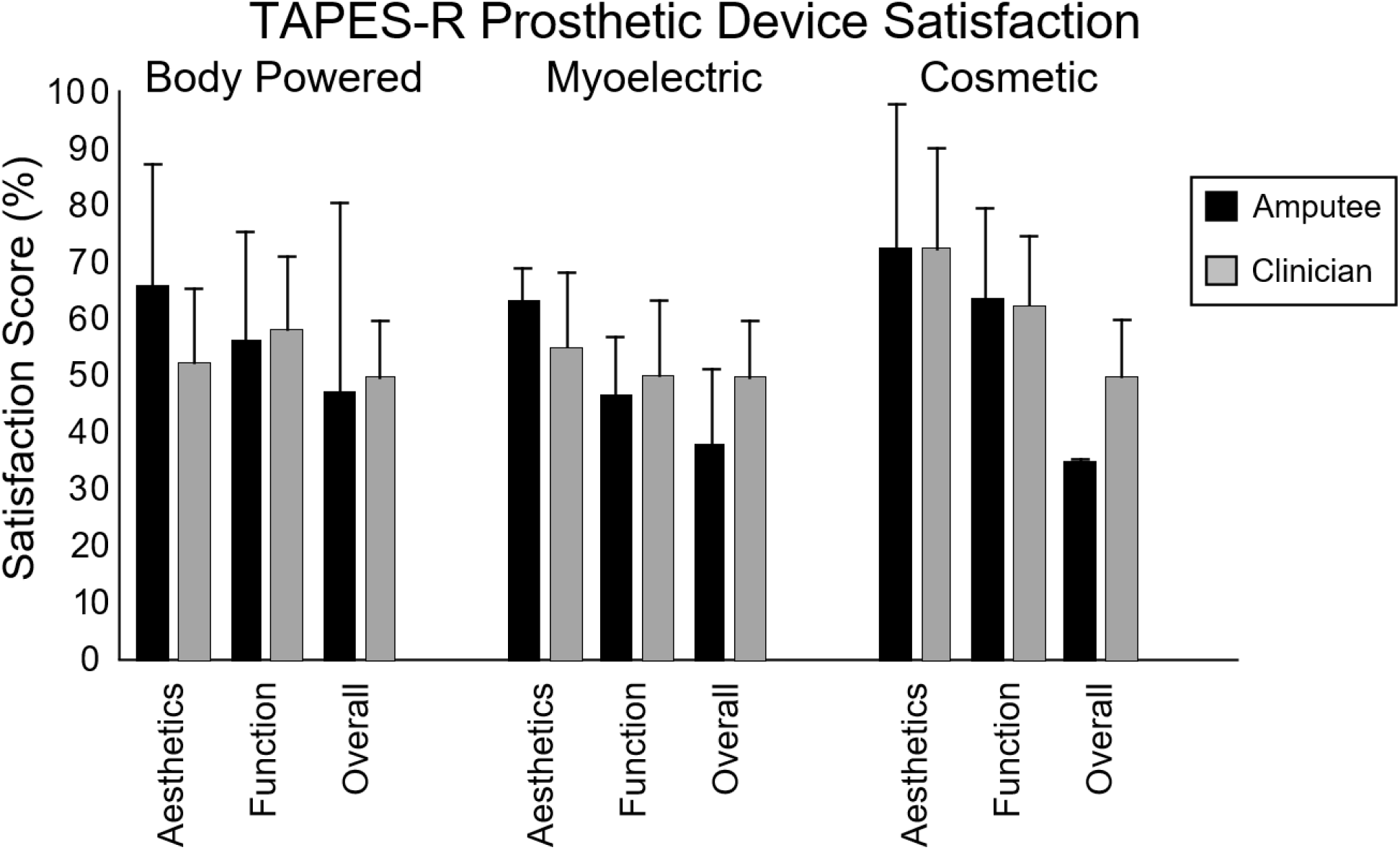
Normalized amputee and clinician ratings of satisfaction with existing prosthetic devices in terms of how well they meet user needs based on device type (Body Powered: clinicians n = 35, amputees n = 10; Myoelectric: clinicians n = 35, amputees n = 7; Cosmetic: clinicians n = 35, amputees n = 2).

#### Stakeholder priorities for prosthetic technology

The amputee and clinician groups completed a free response question to name activities they would like to be able perform with an upper extremity prosthesis. Responses from the amputee group fell into six main categories: activities of daily living (ADLs) (including eating, bathing, dressing, toileting, transferring, and/or continence), recreational activities, manipulation tasks, carrying/grasping tasks, and household chores. Ten out of 18 respondents wrote about ADLs, with common mention of activities relating to eating, dressing, and bathing. Nine out of 18 respondents discussed recreational activities such as playing sports and instruments, and riding motorcycles. Seven respondents desired to better be able to perform manipulation tasks, specifically fine-motor manipulative activities, such as shuffling playing cards and turning the pages in a book. Six respondents sought to be able to perform carrying and grasping activities that require strength and force control, such as bimanual carrying and securing an object in place while manipulating it with the other arm. Three responders wrote about household chores such as mowing the lawn, hammering, and drilling.

Clinician responses about activities that their clients would like to be able to perform covered a broader range of activities as compared to amputee group. Therefore, responses were categorized into ADLs, recreational activities, manipulation tasks, family interaction, and design characteristics (robustness, sensory feedback, intuitive and realistic motion, and force control). Most clinicians (30 out of 32 responses) mentioned ADLs as being important and many also identified recreational activities and hobbies (n=10) and manipulation tasks (n=14). Three clinicians mentioned the ability to hug or hold hands with children and loved ones. Eighteen out of 32 clinicians expressed the desire for improved design characteristics in terms of intuitive and realistic controls (n=7), force control abilities (n=4), improved device robustness to water or extreme temperatures (n=4), and sensory feedback (n=3).

Amputees and clinicians were asked to rate the importance of various hand and wrist motions with an upper extremity prosthetic device. As shown in Figure 2, all proposed functions were rated as being important, however simultaneous flexion/extension of all fingers as a group was the greatest priority for amputees, with the highest ratings of “extremely important”. Clinicians placed the greatest importance (when considering “very important” and “extremely important” ratings) on wrist supination and pronation, followed by finger flexion and extension as a group, and wrist flexion and extension. These two wrist motions were also reported as “very important” or “extremely important” by the majority of amputees. Three quarters of clinicians also reported thumb abduction and adduction abilities for prosthetic devices as very or extremely important.

**Figure 2.**
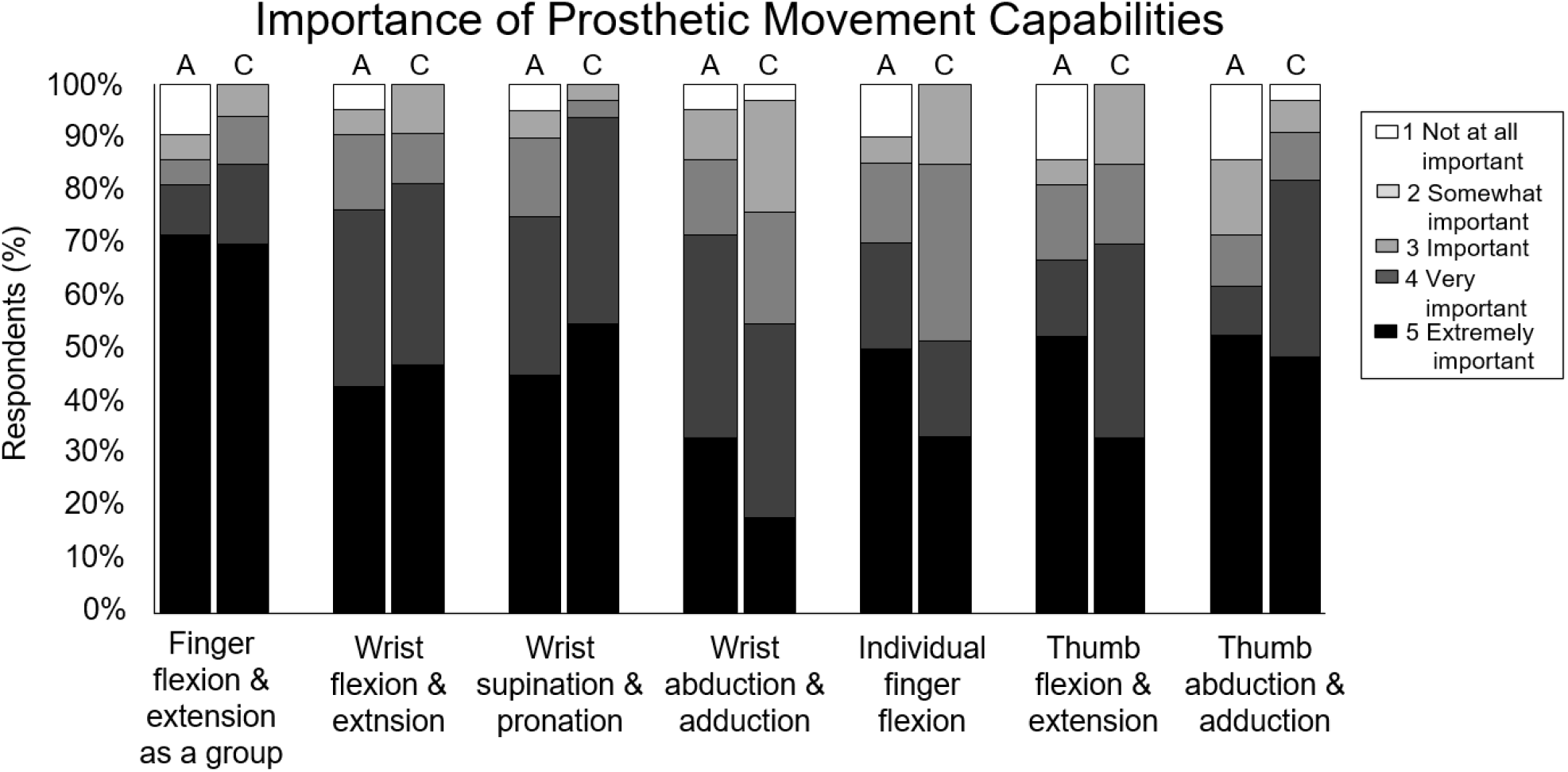
Ratings of importance of prosthetic movement capabilities for amputees (A) and clinicians (C) on a scale from 1 (white) to 5 (black) where 1 is “Not at all important” and 5 is “Extremely important”. Movements are ordered from most to least important based on the amputee group ratings of “very important” and “extremely important”.

#### Technology Acceptance

Amputees, clinicians, and regulators were asked to rate the importance of seven criteria individuals considered when choosing to purchase or adopt technological devices (Figure 3). The top three priorities, considering ratings of “very important” and “extremely important”, for amputees were ease of use, meeting the needs of the user, and cost, respectively. Clinicians and regulators placed the highest importance on meeting the needs of the user, also emphasizing safety, cost, and ease of use among their top priorities. Less than half of amputees reported privacy as an important factor in choosing technology, while two-thirds of regulators felt this was a very important or extremely important factor. All three stakeholder groups placed low importance on the attractiveness and visibility of the technology. Amputees, clinicians, and regulators were also surveyed about their beliefs about technology in general (Appendix B). Technology acceptance scores were generally consistent across groups (H = 2.4, p = 0.31) with amputees reporting 71.5% ± 11.4% acceptance, clinicians 67.6% ± 7.4%, and regulators 65.3% ± 10.1% on a 0 (not at all accepting) to 100% (completely accepting) scale.

**Figure 3.**
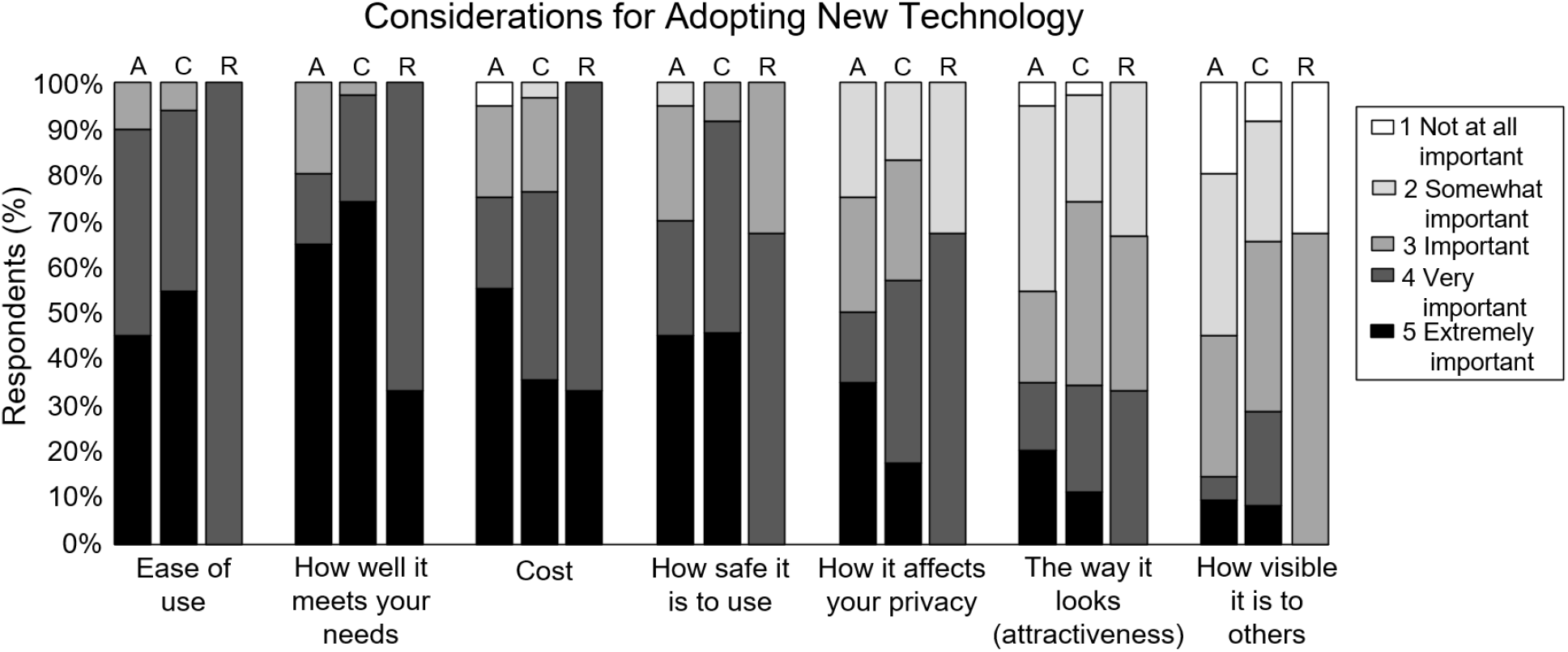
Ratings of important considerations for adoption of new technology among amputees, clinicians, and regulators on a scale of 1 (white) to 5 (black) where 1 is “Not at all important” and 5 is “Extremely important”. Presented in the order amputee (A), clinician (C), regulator (R). Items ordered on amputee ratings from most to least important.

#### Acceptance of a Proposed Advanced Sensorimotor Prosthesis

Participants from all three stakeholder groups were asked to list up to three “likes” and “dislikes” about the proposed implanted sensorimotor prosthesis following the video introduction (Supplementary Video 1). Most participants expressed the device-enabled sensory feedback as one of their likes (13/19 amputees, 29/33 clinicians, and 3/3 regulators). Fourteen out of 19 amputees, 23 out of 33 clinicians, and 1 out of 3 regulators liked that the hypothetical prosthesis would enable fine motor control of the fingers. Other common likes were the innovative design (3/19 amputees, 2/3 regulators) and the perceived ease of use (7/33 clinicians, 1/3 regulators). Clinicians and regulators were most likely to note appreciation for the device’s fully implanted design (11/33 clinicians, 2/19 amputees, 2/3 regulators commented on this feature). Conversely, nine out of 18 amputees, 29 out of 32 clinicians, and 2 out of 3 regulators expressed concern over the spinal cord stimulation and surgical risks associated with the implanted sensorimotor prosthesis. Seven individuals had amputations at or above the elbow and would therefore be unable to use the control scheme as described (implanted electrodes in the forearm); four of these individuals disliked the limited target population for this device. Five of the clinicians also felt this was a limitation of the proposed prosthesis. Other clinician concerns pertained to funding difficulties (n=9) and device reliability (n=8), weight (n=6), and complexity (n=4). Amputee dislikes included the durability and reliability of the device (n=5 out of 18), the non-realistic appearance (n=2), and possible injury to the residual limb due to surgical implantation of the electrodes (n=2).

Eleven out of 21 amputees who completed the Proposed Technology part of the survey reported they would choose to use the proposed sensorimotor prosthesis; seven of these individuals had amputations at or above the elbow. Twelve out of 35 clinicians reported they would recommend the device to their patients. The three stakeholder groups were asked a series of questions to gauge their acceptance of the proposed device (Appendix C) and a normalized acceptance score (0%=no acceptance, 100%=complete acceptance) was computed. Amputees reported a significantly higher level of acceptance (75.5% ± 10.0%) than clinicians (68.8% ± 6.5%; U = 171.5, p = 0.005) but not regulators (67.8% ± 4.6%; U = 14.0, p = 0.166). Further, amputees who experienced residual limb pain expressed significantly greater acceptance of the device (79.4% ± 5.3%, n = 10) compared to those who did not experience residual limb pain (72.3% ± 7.5%, n = 10) (U = 21.0, p = 0.03). No significant difference in acceptance was observed between individuals with phantom limb pain (n=10) and those without (n=11) (U = 26.5, p = 0.08). We noted a trend that higher satisfaction with current prosthetic technology correlated to lower acceptance of the proposed technology though this did not reach statistical significance (rho = -0.479, p = 0.083). Surprisingly, amputation level was not related to proposed device acceptance (U = 43.0, p = 0.61 when stratified by below elbow (n=11) vs. at or above elbow (n=10)).

#### Preferences Related to the Proposed Advanced Sensorimotor Neuroprosthesis

Analysis of the responses to each question that was used to compute the overall acceptance score provide a better understanding of prosthesis characteristics that drive the stakeholders’ acceptance of the proposed device as a whole (Fig. 4). Ratings of characteristics with a positive connotation revealed stakeholders’ preferences of the sensory capabilities and complex hand movements afforded by the proposed sensorimotor prosthesis. All stakeholder groups agreed that it was important to develop an advanced sensorimotor prosthesis like the theoretical one from the video (Supplementary Video 1), while amputees and clinicians felt more strongly about the government investing resources into device development than regulators did. All groups agreed, to varying degrees, that the device should be able to work with many prosthetic hands. More than half of amputees and clinicians agreed or strongly agreed that a device like the proposed sensorimotor prosthesis would make people’s lives easier. However, the majority of stakeholders did not feel as though the implanted sensorimotor prosthesis would be easy to learn to use.

**Figure 4.**
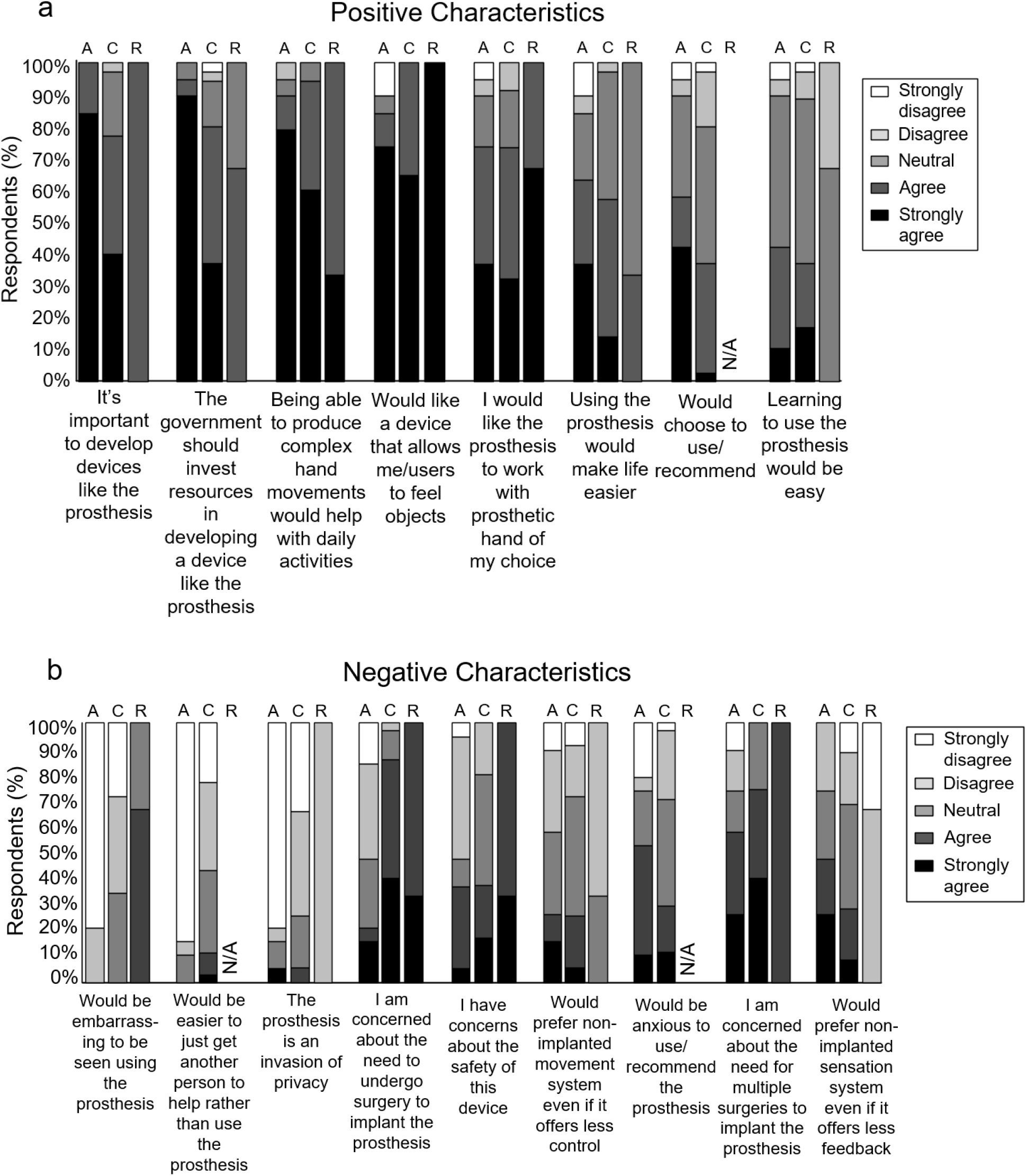
Ratings of acceptance of the advanced implanted sensorimotor neuroprosthesis among amputees (A), clinicians (C), and regulators (R). Responses for a) positive characteristics and b) negative characteristics recorded on a scale from “Strongly disagree” (white) to “Strongly agree” (black). Higher levels of agreement with positive characteristics (greater representation with darkly-shaded bars) is more supportive of the MyoTouch technology. Higher levels of disagreement with negative characteristics (greater representation with lightly-shaded bars) is more supportive of the MyoTouch technology. Items have been ordered based on amputee agreement from most to least supportive of the neuroprosthetic device.

One fifth of amputees (21%) were concerned about the need to undergo surgery, however nearly all clinicians (86%) and regulators (100%) reported this concern. Similar trends were observed related to the prospect of multiple surgeries occurring for device implantation, one for implantation of the myoelectric control components and one for the spinal cord stimulation components for sensory feedback. Half of amputees and a quarter of clinicians expressed a preference for non-implanted sensory feedback and/or movement control systems even at the expense of the quality of those systems. Just over half of amputees (57%) reported anxiousness with using the implanted sensorimotor device but less than a third of clinicians (28%) were anxious about recommending such a device to their patients. Very few stakeholders expressed a preference for getting help from another person over using the proposed device. Similarly, few stakeholders felt as though the implanted sensorimotor prosthesis was an invasion of user privacy. Neither amputees nor clinicians felt that it would be embarrassing to be seen using the device.

Stakeholders were also asked about acceptable design characteristics for implanted sensorimotor prosthesis calibration, reliability, and battery life (Table 2). Calibration of such a system may be needed to map muscle activity into control signals or to establish stimulation parameters for sensory feedback. The majority of stakeholders thought it was acceptable to perform a short daily calibration. Two out of twenty amputees preferred calibration only be performed once per week, with eight clinicians and one regulator also finding this frequency acceptable. Only two amputees and three clinicians desired very infrequent (i.e., once per month) calibration.

**Table 2.** Desirable design characteristics for sensorimotor neuroprosthetic technology.

|  |  | **Amputee (n=20)** | **Clinician (n=35)** | **Regulator (n=3)** |
| --- | --- | --- | --- | --- |
| **Calibration** | **1 time / month** | 2 (10.0%) | 3 (8.6%) | 0 (0.0%) |
|  | **1 time / week** | 2 (10.0%) | 8 (22.9%) | 1 (33.3%) |
|  | **1 minute / day** | 4 (20.0%) | 11 (31.4%) | 1 (33.3%) |
|  | **5 minutes / day** | 7 (35.0%) | 11 (31.4%) | 1 (33.3%) |
|  | **5 minutes / time someone puts it on** | 5 (25.0%) | 2 (5.7%) | 0 (0.0%) |
| **Reliability** | **100%** | 8 (40.0%) | 11 (32.4%)<sup>†</sup> | 0 (0.0%) |
|  | **99%** | 2 (10.0%) | 7 (20.6%)<sup>†</sup> | 0 (0.0%) |
|  | **95%** | 4 (20.0%) | 5 (14.7%)<sup>†</sup> | 2 (66.7%) |
|  | **90%** | 3 (15.0%) | 6 (17.6%)<sup>†</sup> | 0 (0.0%) |
|  | **>80%** | 3 (15.0%) | 5 (14.7%)<sup>†</sup> | 1 (33.3%) |
| **Battery life** | **Longer than 1 day** | 2 (10.0%) | 8 (22.9%) | 0 (0.0%) |
|  | **1 day** | 5 (25.0%) | 17 (48.6%) | 2 (66.7%) |
|  | **8 hours** | 8 (40.0%) | 4 (11.4%) | 1 (33.3%) |
|  | **4 hours** | 4 (20.0%) | 5 (14.3%) | 0 (0.0%) |
|  | **1 hour** | 1 (5.0%) | 1 (2.9%) | 0 (0.0%) |

<sup>†</sup>n=34 clinicians answered this question

In general, reliability expectations were very high among all three groups. Half of the amputee and clinician groups reported that the implanted sensorimotor prosthesis should be 99% or 100% reliable. At least half of clinicians and regulators reported the battery would need last a full day or even longer. Approximately one third of amputees had this same requirement for battery life, however most (65%) stated that 8 hours or less of battery life would be sufficient. All groups felt the cost of the neuroprosthetic device should be covered by insurance.

All three regulators agreed that the device should be covered by insurance. One regulator noted that this device would likely require a new medical code in order to be reimbursed. When asked about the proposed device’s potential for reimbursement, two of the three regulators emphasized the importance of evidence supporting the device’s ability to provide a meaningful benefit for the user, and that these benefits must exceed the risks associated with the device for it to receive approval for reimbursement. The third regulator emphasized the value of reliability of the device over time.

## Discussion

### Summary of findings

This study sought to understand stakeholder perspectives broadly as they relate to the impact of amputation, existing prosthetic devices, and emerging advanced technologies. Importantly, in addition to trying to comprehensively understand priorities related to amputation and prosthetics, we compare the perspectives of amputees to those of the clinicians who are prescribing this technology. First, we found that amputees were more accepting of a proposed advanced sensorimotor neuroprosthesis than clinicians, however they also expressed greater anxiety about using the device, emphasizing the importance of shared decision-making when it comes to device selection ^[43]^. The majority of amputees and clinicians agreed that the proposed sensorimotor device would make people’s lives easier (Fig. 4a). Second, we found that amputees reported clinically significant functional impairment as measured by the DASH and over one-third of prosthesis users were still limited by needing help from another person. Finally, we found the majority of stakeholders prioritized ADL performance improvements and user safety for future upper extremity device design. Incorporation of stakeholder preferences can inform the design process with the goal of developing devices with higher rates of device utilization and satisfaction than current technology. Furthermore, the inclusion of regulators in the design and evaluation of upper extremity prosthetic technologies provides insight into potential regulatory barriers and may help avoid unnecessary challenges down the road, especially given the importance of cost in technology decision-making (Fig. 3).

#### Impact of amputation and satisfaction with existing technology

Consistent with current literature ^[7, 55]^, amputee data revealed poor integration of their prosthesis (or residual limb) into performance of activities of daily living, regardless of prosthesis adoption. Twenty out of 21 amputees utilize their sound limb for all activities they are able to perform. Amputee and clinician reports of satisfaction with existing upper extremity prosthetic technology were similar although amputees generally reported slightly less overall satisfaction with their devices. Satisfaction with the functionality and aesthetics of one’s current prosthesis were highest for cosmetic prostheses, followed by body-powered prosthesis users, then myoelectric device users (Fig. 1). Biddis and Chau also found low ratings of satisfaction among users of body-powered prostheses, however their review found that users of cosmetic prostheses were similarly dissatisfied ^[7]^.

#### Stakeholder priorities for prosthetic technology

A 2016 review by Cordella, et al., suggested that user satisfaction would be improved with prosthetics that improve performance of ADLs, integrate sensory feedback, and use advanced motor control schemes ^[35]^. More recently, Zheng, et al., found prosthetic users value device durability, aesthetics, and ability to provide the user with increased dexterity ^[38]^. User- and clinician-reported priorities for improvement in the present study aligned with these priorities since they centred around improved ability to perform ADLs, hobbies, and object manipulation. Further, high levels of agreement were reported across all three stakeholder groups relative to the statement “I would like a prosthesis that allows users to feel objects” (Fig. 4a). Similarly, Biddiss, et al., found that users of electric upper-limb prostheses valued development of devices with sensory feedback and improved dexterity ^[34]^. Interestingly, Engdahl, et al., found users had greater interest in a prosthetic device performing basic features “like opening and closing the hand slowly” over more complex control features such as those desired by participants in the present analysis ^[36]^.

High ratings of importance for simultaneous finger flexion/extension and increased wrist range of motion capabilities were consistent with reported desired design parameters from other studies, as well as the clinician reports of high importance for thumb abduction/adduction ^[35, 36]^. Though specific priorities for wrist motions are not found in the literature, Resnik, et al., found the DEKA arm, which offers powered two-degree-of-freedom wrist motion, has been met with high ratings of satisfaction among users and clinicians ^[14]^. The recognized importance of wrist positioning is likely related to the increased capabilities of object interaction and manipulation it allows. Overall, stakeholders reported high levels of importance for all types of wrist and hand movements surveyed in the present analysis indicating a desire for a multi-functional device with a wide range of manipulation capabilities.

#### Preferences Related to the Proposed Advanced Sensorimotor Neuroprosthesis

Amputees reported significantly greater acceptance of the proposed implantable prosthetic technology than clinicians (75.5% vs. 68.8% acceptance). This does not appear to be due to a bias towards technology acceptance preferences in general, which were similar across all three groups (71.5% for amputees, 67.6% for clinicians, 65.3% for regulators). Concerns about the proposed device included surgical risk, questions about robustness, and potential damage to the residual limb during electrode implantation. Surveys of adults with upper-limb amputations have shown differing perspectives on their willingness to undergo surgery to implant electrodes for prosthetic control. Typically, younger amputees with acquired unilateral limb loss have been most interested in invasive devices ^[42, 43]^. Surgery-related concerns were more prevalent among clinicians and regulators than amputees in our study (Fig. 4b). Consistent with expectations ^[56]^, greater consideration among all stakeholders was given to user-centred outcomes such as meeting the user’s needs and the safety of the device (Fig. 3). Of note, few stakeholders noted the fully implanted design as a feature of the device that they liked, indicating that this may not be a priority.

Reliability requirements for emerging prosthetic technology were high across all groups. Amputees were more accepting of more frequent calibration or lower battery life than clinicians or regulators. This prioritizing of reliability in a proposed upper-extremity prosthesis is consistent with Janssen’s findings in amputees and clinicians/researchers ^[56]^. These ratings can help direct engineering focus as designs are optimized, however more work is needed to understand motivations behind their ratings.

#### Limitations

While this study was able to include the important perspectives of device regulators, the small sample size and limited experience of this group limited our ability to robustly compare perspectives to the amputee or clinician groups. In addition, amputee group sizes were small when comparisons by prosthesis type were considered (Figure 1), likely contributing to the lack of statistical significance for these analyses. Further, the rate of prosthesis use among the amputees surveyed in this study (81%) was high as compared to general device use that has been previously reported in the United States (55-65%) ^[5, 7]^. Therefore, our results may not generalize to less active prosthesis users.

It is important to mention that the fully implanted sensorimotor neuroprosthesis presented (called the “MyoTouch” for brevity in the surveys) is a hypothetical device that was proposed as an example of an advanced prosthesis that combines implanted myoelectric control and somatosensory feedback. While our findings may be broadly applicable to implantable neuroprosthetics, we caution that the results are likely impacted by the specifics of this conceptual device. Interestingly, acceptance of this technology was not impacted by level of amputation (below elbow vs. at or above elbow) even though the proposed sensorimotor device was presented as targeting those with amputations below the elbow. This may indicate good translatability of the results to neuroprosthetic technology in general.

Finally, the TAPES-R questionnaire reported in the analysis here was adapted to be more prosthesis-focused instead of amputation-focused. Therefore, it is not appropriate to directly compare the TAPES-R results here to other studies.

## Conclusions

The present study surveyed amputees, clinicians, and regulators on satisfaction with existing prosthetic technology and their expectations for future devices. Stakeholders were accepting of the advanced functionality of emerging neuroprosthetic technology and reported high expectations for reliability of this type of device. Concerns about surgical risk were common, particularly among clinicians and regulators. In general, amputees were more tolerant of risk and accepting of the proposed technology than the other groups. Overall, their priorities focused on improving the abilities of upper limb prosthesis users while maintaining their safety; stakeholder perspectives should be considered as advanced neuroprosthetic technology is developed to ensure that it meets users-identified needs and expectations.

## Data Availability

Data available upon request.

## Acknowledgements

The authors would like thank Santiago Munoz for contributions to the survey design and Debbie Harrington for coordinating study activities and for assistance with data collection.

## Appendix A: Disability of the Arm, Shoulder, and Hand – limb use questionnaire

For the activities listed in the previous question, please mark which arm you use most often to complete the tasks: intact arm, arm with amputation, or both.

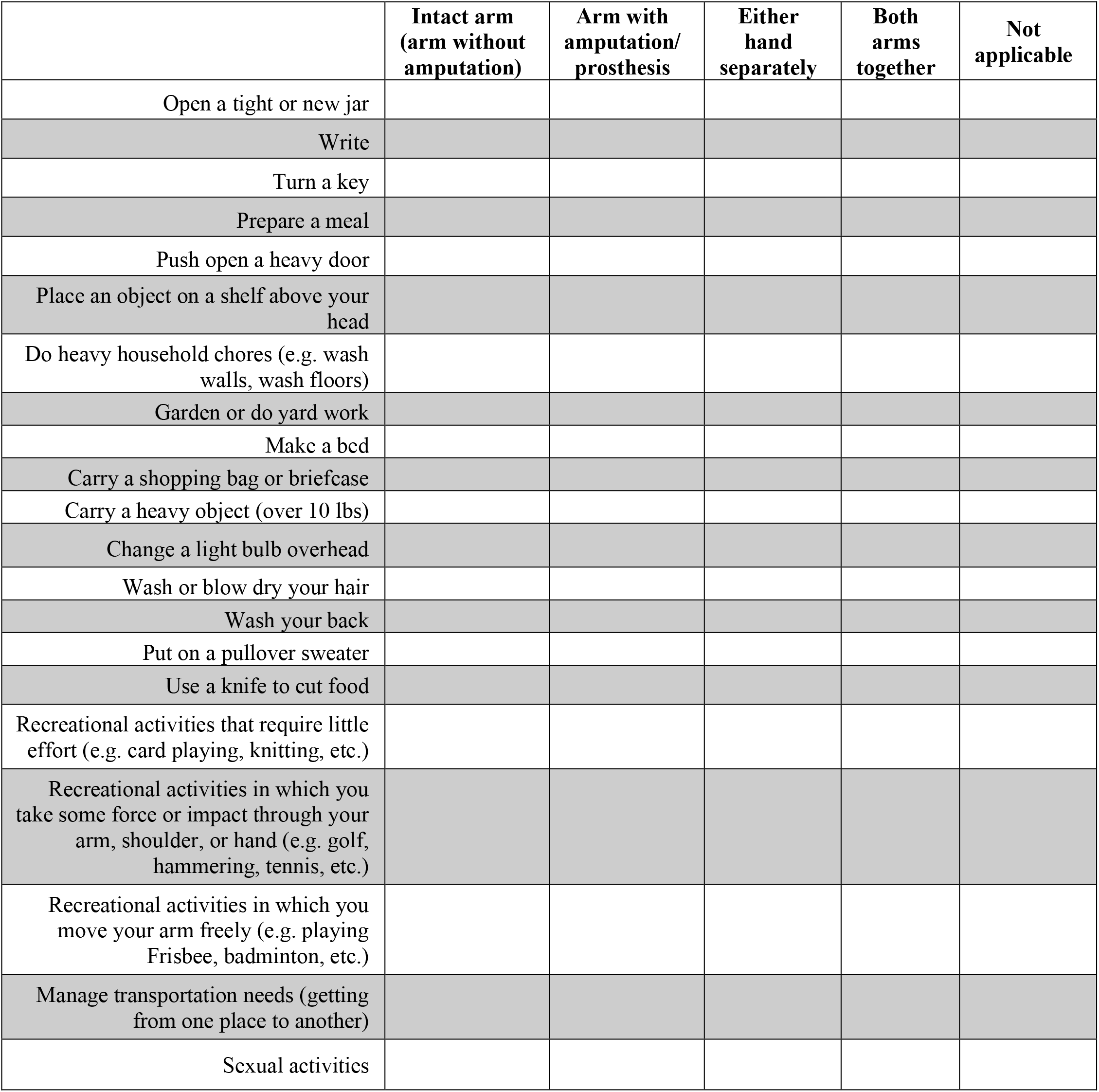

## Appendix B: Technology acceptance survey

Responses from parts 1 and 2 were combined to come up with a “Technology acceptance score” presented as a percentage where a higher score indicates better acceptance of technology. Shaded components were considered positive characteristics (scored 1-5 where 5 is “Completely” agreeing with the statement) and unshaded were considered negative characteristics (scored 5-1 where 1 is “Completely” agreeing with the statement).

1. In general, to what extent do you believe that technology:

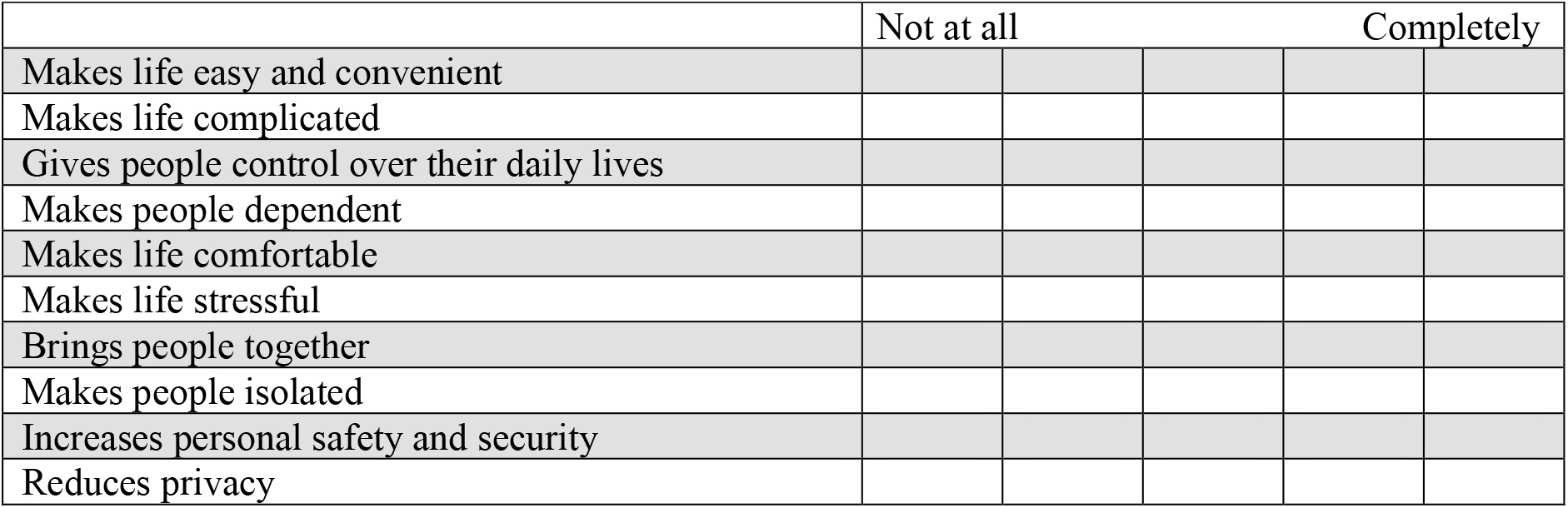
2. How accurately does each of the following phrases describe you?

3. How important is each of these factors for you when choosing technology?*

2. How accurately does each of the following phrases describe you?
|  | Not at all |  |  |  |  | Completely |
| --- | --- | --- | --- | --- | --- | --- |
| I like to keep up with the latest technology |  |  |  |  |  |  |
| I generally wait to adopt new technology until all the bugs have been worked out |  |  |  |  |  |  |
| I enjoy the challenge of figuring out high tech gadgets |  |  |  |  |  |  |
| I feel confident that I have the ability to learn to use technology |  |  |  |  |  |  |
| Technology makes me nervous |  |  |  |  |  |  |
| If a human can complete a task as well as technology, I prefer to interact with a person |  |  |  |  |  |  |
| I like the idea of using technology to reduce my dependence on people |  |  |  |  |  |  |

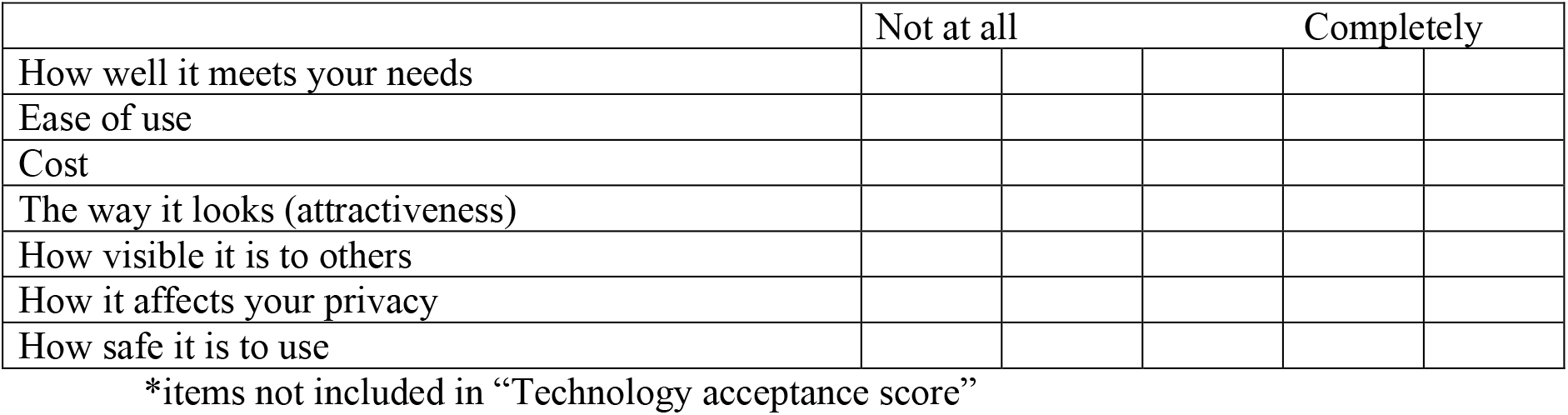

## Appendix C: MyoTouch Acceptance Questionnaire

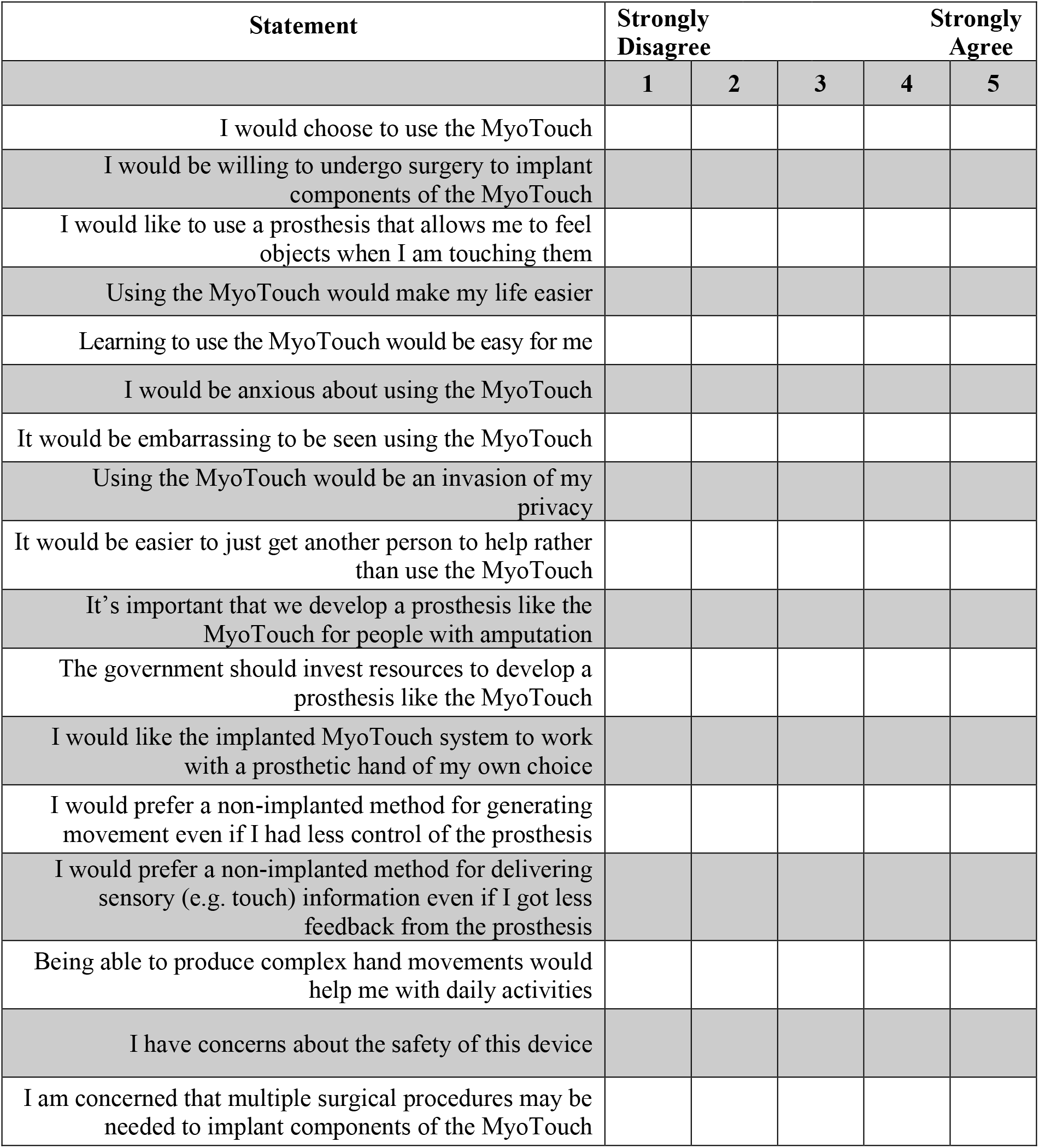

